# Kidney Dialysis Service: Experiences of Consumer and Provider and Quality of Life of End Stage Kidney Disease Patients in Parbat, Nepal

**DOI:** 10.1101/2024.10.05.24314694

**Authors:** Mamta Dhakal, Prabin Sharma, Anjali P.C., Suman Sharma

## Abstract

**Background/Objectives:** The kidney dialysis patients and service providers possess a wide range of experience relating to the service, and quality of life is an important measure to evaluate holistic approach to chronic disease management such as kidney disease. This study was conducted to find out the experience of kidney dialysis patients and service providers and quality of life of end stage kidney disease patients.

**Methods:** A qualitative study was conducted, from March 2024 to August 2024, among 20 end stage kidney disease patients visiting Parbat hospital for dialysis and 7 dialysis service providers, doctor and nurses, using in-depth interview guidelines, kidney disease quality of life-36 instrument (for end stage kidney disease patients) and job satisfaction survey tool (for service providers). Thematic analysis was performed from the transcribed and translated recorded information to carry out the actual theme.

**Result:** Nine (9) themes were extracted for experiences of kidney dialysis patients. They were (i) perception on end-stage kidney disease, (ii) perception on free kidney dialysis service at the hospital, (iii) administrative process for free dialysis service, (iv) feasibility with the schedules of dialysis session, (v) quality of care and response of service providers (vi) duration of dialysis session (vii) desires and challenges towards kidney transplantation, (viii) on recommending the dialysis site and (ix) further expectations for improvement. The quality of life of end-stage kidney disease patients was found to be moderate. Similarly, nine (9) themes were extracted for experiences of service providers. They were (i) patient personalized care and understanding patients’ behavior (ii) addressing queries and concerns of the patients (iii) rewarding aspect of the job (iv) learning acceptance and witnessing positivity (v) availability of safety measures and protective protocols (vi) training and professional development experience (vii) reflection on job security (viii) challenging aspect of the job (ix) further expectations and requests. The kidney dialysis service providers were found to be ambivalent, neither satisfied nor dissatisfied with the job.

**Conclusion:** Patients share an overall good experience of free kidney dialysis service at Parbat hospital while the service providers also have a positive experience working in the dialysis section. However, expectations for provision of free medications and availability of nephrologist by the end stage kidney disease patients direct towards the reforms for improved service delivery. The doctor and nurses should be provided with more opportunities for professional development and safety should be a priority. These respective vigilance and actions for, shall address the quality of life and job satisfaction as well.

## Introduction

The last permanent stage of chronic kidney disease when the kidney/s do not function on their own and need dialysis or transplantation is referred to as end stage kidney disease (ESKD).(1) Dialysis is a process to remove the waste products and fluid in the body, which when left untreated can lead to unpleasant symptoms and be fatal as well, from the blood when the kidneys stop to work properly.(2)

The incidence of end stage kidney disease has plateaued or declined in many countries, nevertheless, the prevalence has rose by a median of 43% from 2003 to 2016.(3) It is estimated that the incidence of end stage kidney disease in Nepal to be 2900 patients (100 per million population).(4)

Globally, kidney dialysis services are predominant therapy in managing ESKD.(3) The Government of Nepal has launched and implemented free dialysis services from 2016 to 1975 patients through 42 dialysis centers.(4)

Quality of life (QoL), according to the World Health Organization is “an individual’s perception of their position in life in the context of culture and value systems in which they live and in relation to their goals, expectations, standards and concerns”.(5) Quality of life is an important measure to evaluate holistic approach to chronic disease management such as chronic kidney disease and patient outcomes as well as quality of care.(6,7) The quality of life of the patients undergoing kidney dialysis therapy is low.(6)

The kidney dialysis patients perceive that dialysis prolongs life, it is indispensable part of their life, clears the bodily toxins, makes them feel better, while family financial support, physical limitation and emotional distress are the challenges.(8) The health service providers working in the kidney dialysis are usually satisfied with their responsibilities and duties.(6) However, kidney dialysis service providers have low personal accomplishment and are dissatisfied with certain job areas.(8) The kidney dialysis workforce is at critical point as well, reporting burnout.(9)

Kidney diseases are often diagnosed late and the global disease burden is underappreciated. By 2040, chronic kidney disease is estimated to be the fifth leading cause of mortality globally.(6) It was the tenth leading cause of death in Nepal in 2019. (8)

The quality of life (QoL) of patients is increasingly becoming important in the delivery of health service including kidney disease. Evaluating QoL in the context of kidney dialysis can help identify specific areas where interventions are needed to enhance overall patient well-being.(10) However, there are limited evidences relating to QoL, particularly among the kidney patients in stage 5 – end stage.(11)

The assessment of experiences of kidney dialysis patients can shed light on the psychological, social and economic burdens they go through. Likely, the insights of health service providers can highlight the operational challenges and systematic issues within the healthcare infrastructure. These perspectives are essential for developing patient-centered care models and policy interventions aimed at improving the quality of life for end stage kidney disease patients. The experiences of both service consumers and providers in the context of dialysis services are crucial. These experiences can provide insights into the dialysis services, essential for improving patient outcomes and enhancing service delivery.

Parbat hospital has launched kidney dialysis service from 2019 which has now been extended in 2024 to a new infrastructure. Thus, to address the existing scenario in Parbat district in kidney dialysis service, this study was conducted with the aim to assess the experiences of kidney dialysis patients and service providers, doctor and nurses, and quality of life of end stage kidney disease patients of Parbat district, Nepal.

## Methods

### Study design, settings and population

This was a qualitative study conducted among end stage kidney disease patients visiting Parbat Hospital for dialysis and dialysis service providers, doctor and nurses, working in the kidney dialysis section of Parbat Hospital. Parbat hospital has launched kidney dialysis service from 2019 which has now been extended in 2024 to a new infrastructure providing dialysis service to 21 kidney disease patients. The study period was from March 2024 to August 2024. 15 kidney dialysis patients and 5 service providers were interviewed to assess the experiences and was stopped once data saturation was attained. However, along and beyond the interview, in total, 20 kidney dialysis patients and 7 service providers were respectively interviewed to assess the quality of life and job satisfaction. One (1) participant was unavailable at the time of data collection since s/he has gone for follow-up outside the district. Healthcare providers directly involved in the care of ESKD patients undergoing dialysis and kidney dialysis patients who are residents of Parbat district, Nepal, or those who receive dialysis services in Parbat were included in the study.

### Data collection techniques and tools

The data collection technique was face-to-face interview. The study tools for kidney dialysis patients were In-depth Interview Schedule and Kidney Disease Quality of Life (KDQOL-36) instrument. The KDQOL-36 is a free-access instrument specifically designed for kidney disease patients.(12) The study tool for service providers were In-depth Interview Schedule and Job Satisfaction Survey (JSS) 36-item tool.(13) The in-depth interview schedules were developed after extensive review of related literature consists of major questions along with probing questions related to experiences separately for both dialysis patients and service providers.

### Data processing, management and analysis

The scores of tool to assess quality of life of the kidney disease patients were entered and analyzed in a stand-alone Microsoft Excel scoring tool developed by KDQOL working group.(12) The scores of job satisfaction survey (JSS) tool were entered and analyzed in Microsoft Excel as per the instructions for scoring the tool.(13)

The recordings of the in-depth interview were listened properly and transcribed into words (to English and full text). The transcribed information was categorized in different categories. During coding of the categories, each transcript was reviewed and codes were be assigned. The coded text was then be sorted in one group to organize the information to make easier for analysis and the content was reduced to create a meaningful text to reflect the original view. Lastly, the actual theme was carried out from the transcript.

### Strengths and limitations of the study

This study has several strengths as it is one of the few studies assessing the experiences of end stage kidney disease patients in Nepal. The study was hospital based study. For this reason the chronic kidney disease patients of the community other than those visiting Parbat Hospital could not be assessed. Similarly, experiences of patients in Parbat district might not be generalizable to other regions with different healthcare infrastructure, socioeconomic conditions, or cultural contexts.

## Results

### Background information of kidney dialysis patients

Table 1 describes the background information of the participants, which includes information on age, sex, ethnicity, marital status, family type, education level, occupational status and income status of participants. The age of participants ranged from 24 to 77 years.

**Table 1.** Background characteristics of kidney dialysis patients n=20.

| **Characteristics** | **Number** | **Percentage** |
| --- | --- | --- |
| **Sex** |  |  |
| Male | 14 | 70 |
| Female | 6 | 30 |
| **Age** |  |  |
| <40 | 4 | 20 |
| 40-49 | 4 | 20 |
| 50-59 | 7 | 35 |
| >59 | 5 | 25 |
| **Ethnicity** |  |  |
| Dalit | 10 | 50 |
| Upper caste | 8 | 40 |
| Disadvantaged Janajatis | 2 | 10 |
| **Marital Status** |  |  |
| Married | 17 | 85 |
| Separated | 1 | 5 |
| Widow/Widower | 1 | 5 |
| Unmarried | 1 | 5 |
| **Family Type** |  |  |
| Joint | 11 | 55 |
| Nuclear | 9 | 45 |
| **Education** |  |  |
| Illiterate | 2 | 10 |
| Informal | 5 | 25 |
| Primary level | 6 | 30 |
| Secondary level | 6 | 30 |
| Bachelor and above | 1 | 5 |
| **Occupation** |  |  |
| Agriculture | 10 | 50 |
| Home-maker | 4 | 20 |
| Unemployed | 4 | 20 |
| Retired with pension/gratuity | 1 | 5 |
| Service (Private/Government) | 1 | 5 |
| **Income status** |  |  |
| Adequate | 11 | 55 |
| Inadequate | 9 | 45 |

### Kidney disease related information of dialysis patients

Table 2 presents the information regarding kidney disease characteristic of dialysis patients including length of diagnosis, dialysis and dialysis at Parbat Hospital, health insurance type cause of kidney disease and desire for transplantation.

**Table 2.** Kidney disease related characteristics of dialysis patients n=20.

| **Characteristics** | **Number** | **Percentage** |
| --- | --- | --- |
| **Length of diagnosis of kidney disease** |  |  |
| <2years | 3 | 15 |
| 2-5 years | 12 | 60 |
| 6-9years | 3 | 15 |
| >9 years | 2 | 10 |
| Minimum=1year 4months, Maximum=13 years |  |  |
| **Length of dialysis** |  |  |
| <2years | 8 | 40 |
| 2-5 years | 10 | 50 |
| >5 years | 2 | 10 |
| Minimum=1 month, Maximum=7 years |  |  |
| **Length of dialysis in Parbat Hospital** |  |  |
| <2years | 8 | 40 |
| >2 years | 12 | 60 |
| Minimum=1 week, Maximum=4 years |  |  |
| **Health insurance type** |  |  |
| Health insurance of Nepal Government | 20 | 100 |
| **Cause of kidney disease** |  |  |
| High Blood Pressure | 17 | 85 |
| High Blood Pressure and Diabetes | 3 | 15 |
| **Desire for transplantation** |  |  |
| Yes | 10 | 50 |
| No | 10 | 50 |

### Experience of kidney dialysis patients

i. Perception on end-stage kidney disease, Patients expressed feelings of sadness and frustration regarding their condition. They lament their inability to travel, play, and work like their friends, highlighting the social and physical limitations imposed by kidney disease. The mention of attempting to enroll in the Nepal Army but being unable to succeed due to their illness underscores the significant impact that kidney disease has had on their aspirations and life goals.

> ……I feel bad about my condition. I cannot travel, play and work like my friends. I tried to enroll in Nepal Army but couldn’t succeed because of kidney disease. P2 In contrast, some patients conveyed a sense of acceptance regarding their condition. While they do not expect to regain strength or return to work, they express optimism that dialysis will extend their life.

> ……I don’t expect to be strong and return to work. However, dialysis will lengthen my life duration P6
ii. Perception on free kidney dialysis service at the hospital, Some of the oldest dialysis patient at the hospital, shared their experience since the service’s initiation. They metaphorically describe the arrival of dialysis provided much-needed relief and comfort. They note a significant decrease in expenses compared to previous treatments, emphasizing the importance of having access to medications through health insurance, which they didn’t have before. This reflects a positive change in their healthcare experience, although they acknowledged that new patients may not fully appreciate the improvements made over the years.

> …. I am the oldest dialysis patient of this hospital. I have taken the dialysis service since its initiation here. When dialysis service started here, it felt like coming under a shadow on a sunny day. I had spent large amount before coming here. Comparatively, our expenditure has declined abruptly. We couldn’t get medicines from health insurance before. Now, we have some medicine under the scheme. I can feel the difference which maybe, new patients won’t experience. P1 Patients expressed satisfaction with the government’s provision of dialysis services, stating that it has positively impacted their lifespan. The patients showed gratefulness for the support provided by the government, recognizing its role in enhancing their quality of life.

> ……The service given by government is very good. The dialysis service has lengthened the lifespan of patients like us P9 Patients appreciated the availability of free dialysis and health insurance for medications, which has significantly helped manage their condition. This highlights the financial relief that comes from these services, underscoring their importance in enabling patients to focus on their health without the burden of high medical costs.

> ……The dialysis is free here and there is health insurance for medicine. These have assisted for my condition P6
iii. Administrative process for free dialysis service, Most patients described the administrative procedures for admission into the free dialysis program straightforward and not overly burdensome. This indicates that the hospital’s administrative systems are relatively efficient in facilitating access to the service.

> ……The administrative processes of admission for free dialysis is neither lengthy nor difficult……P1

> ……The administrative process of admission for free dialysis was easy……P8 Although the paperwork and administrative process were not tedious, some patients explained they faced challenges in securing timely dialysis sessions due to a shortage of available beds. This highlights a common issue in resource-limited settings where demand for dialysis often exceeds available capacity.

> ……The paper work and administrative service for admission is not tedious. However, the turn for dialysis in the initial days were not available due to limited dialysis beds……P9
iv. Feasibility with the schedules of dialysis session, Patients explained they find no difficulty in adapting to changes in their dialysis schedule, whether it’s moving between morning and afternoon shifts or arranging accommodations such as staying in hotels when necessary. This suggests a level of flexibility and comfort in managing their treatment, as they have found ways to work around any inconveniences, such as residing in a hotel or living nearby.

> …My dialysis session is usually scheduled for the second shift of the day starting at 1-2 pm. If, sometimes, the session is scheduled for morning shift, I come a day before and reside in hotel. I find no difficulty in this… P1

> ……I live nearby with my family as tenant. It is easy for me to access the hospital… P2

> ……I am fine with the scheduling of dialysis session. I had my sessions in the morning shift before. Later, another patient came and my shift was changed to the day shift P6 For some patients, they expressed the timing of the dialysis session creates difficulties, especially when the session is scheduled later in the day. Long travel times and the need to stay overnight at a hotel add to the burden of receiving treatment, particularly for those living further away.

> ……It takes around 2 hours to reach the hospital. When the session is after 1-2 pm, it is difficult to return back home. Sometimes, we have to stay here in hotel P10
v. Quality of care and response of service providers, Patients expressed contentment with the overall service, noting that the hospital is clean and that everything operates on time. This suggests that the hospital maintains a good level of hygiene and punctuality, which contributes to a positive patient experience.

> ……I am satisfied with the service here. It is clean here. Everything is timely P3 Patients also acknowledged that the services are satisfactory and noted that the dialysis process seems consistent across different locations. The patient appreciated that they do not have to wait long and highlights the prompt availability of dialysis during emergencies, indicating that the hospital is efficient in managing urgent cases.

> ……The services are fine. Maybe this is the process. How would we know about the quality? However, I have seen the same process wherever I have gone. We don’t have to wait long and even in case of emergency dialysis, they get the time instantly P9 Patients appreciated the way nurses respond to patient queries, noting that they provide proper suggestions. This suggests that the nursing staff is knowledgeable and effective in addressing patients’ concerns, contributing to a positive experience.

> ……The nurses’ responses are good. They suggest us properly for our queries… P9 Most of the patients echoed similar sentiments, expressing satisfaction with the hospital staff’s politeness and responsiveness. The patients also emphasized that the staffs answer their questions well, which reinforces the sense of care and support they receive.

> ……The staff are very good here, They are polite, responsive and answer our queries nicely P12
vi. Duration of dialysis session, Patients shared they experienced mixed emotions, noting that they sometimes feel restless towards the end of the session but also feel that the time passes quickly on certain days. This indicates that their experience may vary depending on their mood or physical state during dialysis.

> ……Sometimes, towards the end of dialysis session, I fell restless. And, sometimes, the time flies P4 Patients also reflected a positive outlook on the dialysis process. Unlike other patients who might find the long sessions tedious some patients expressed a sense of relief and satisfaction during dialysis, as they perceive it as a cleansing process for their body. The treatment makes them feel good, both physically and mentally, showing a positive attitude towards managing their condition. This suggests that for some patients, focusing on the health benefits of dialysis can foster a more optimistic experience.

> ……I do not feel bored during the dialysis session. I feel like my body is getting cleaned. I feel good P6 Patients expressed that they often feel bored during the 3-4 hour dialysis sessions and mentioned that they sometimes sleep to pass the time.

> ……I get bored during the dialysis session… P7

> ……The length of dialysis session is around 3-4 hours. I feel bored. Sometimes, I sleep during the time P11
vii. Desires and challenges towards kidney transplantation, Patients appealed to the Nepal Government to amend transplantation laws, advocating for allowing donors who are not limited to family members. This highlights the emotional and logistical difficulties patients face when relying solely on family members for such a critical decision.

> ……We (family) have planned for kidney transplantation. My sibling was the probable donor. Only cross-matching was remaining to finalize the transplantation. However, he hesitated later. I want to request Nepal Government to amend provisions for kidney transplantation and allow donors to be other than family members as well……P2 Patients expressed a strong desire for a kidney transplant, the financial burden of the transplantation process added to the stress. Patients who have young children, worried about their future and believed that a successful transplant would provide them with more time and a better quality of life.

> ……I desire for kidney transplantation. My parent was ready to donate but cross matching could not happen. The process is costly as well. If I could transplant my kidney, I would live more. I have small children, I worry about them……P10
viii. On recommending the dialysis site, Patients shared they have had positive experiences with the dialysis services at Parbat Hospital. They expressed satisfaction with the services and are willing to recommend the hospital to others, including friends, family, and fellow patients. Their appreciation seems to stem from the quality of care provided by the nursing staff and the overall service environment.

> ……I will recommend Parbat hospital for dialysis for patients like me……P2

> ……I like the services here. I will surely recommend kidney patients like me to visit this dialysis center for the service……P11

> ……I will recommend this hospital to take dialysis service. I will to take dialysis service. I will ask my relatives, neighbors, friends, if any day, they are prescribed dialysis, to be admitted here……P8 Contrary, some patients acknowledged the good service, responsive nurses, and positive environment, the absence of a nephrologist (kidney specialist) is a significant issue for them. This lack of specialized medical expertise prevented some patients from recommending the hospital for dialysis, despite the other positive aspects.

> ……I will not recommend this site for dialysis because of lack of specialized doctor only. The nurses are good here, very responsive. The service and environment is good but lack of nephrologist is the issue……P5
ix. Further expectations for improvement Patients appreciated the quality of care provided by nurses and expect the high standard of service to continue. This reflects patient satisfaction with the healthcare staff and the overall treatment process.

> ……The services are good here. The nurses are caring and responsive. I expect the service to continue in the coming days as well……P1 Patients highlighted that the hospital is not equipped to handle critical situations, specifically mentioning the absence of an ICU and a kidney specialist. Some of the patients mentioned that they have only had one discussion with a doctor. Patients emphasized the need for a nephrologist, as critical situations can arise unexpectedly, and having a specialist would help mitigate risks and improve patient outcomes.

> ……When critical situations arise here, the hospital is not equipped for so; example: there is no ICU. The hospital has doctors but do not have specialist doctor for kidney service……P2

> ……The major issue is of doctor. I have discussed with doctor only once. We might have critical situation any time. It would be better and the risk of patient would reduce if a specialized doctor would be available……P5 Patients advocated for the government to provide free kidney transplantation for poor patients who have donors. Though some patients do not currently have a donor, the patient emphasizes that economically disadvantaged patients with donors could benefit from this life-saving surgery, potentially offering them a new lease on life.

> ……The government should make free provisions for kidney transplantation too. Though, I do not have any donor, patients economically poor, with donor, could get a new life…… P9 Patients also mentioned the co-payment requirement for medicines under the Health Insurance scheme, which they feel should be waived for chronic patients like themselves. Similarly, the patients pointed out that while dialysis is free, the cost of medications remains a significant financial strain, especially as they are the sole breadwinner of their family. They suggest that making medicines free for dialysis patients would be a crucial step toward reducing the financial burden.

> ……For the medicine available free from Health Insurance previously, we have to pay 10% co-payment. This system should not be made for chronic patients like us……P4

> ……It would be better if medicines are also free. I am the only breadwinner of the family. Though the dialysis is free, the medicines need to be taken regularly and are costly……P8 While dialysis treatment itself is free, patients explained they still face significant monthly expenses related to travel, food, lodging, and medication, amounting to around 20-25 thousand Nepalese rupees. Patients expressed the need for the Nepal Government to provide free medications to further alleviate the financial burden on patients.

> ……Though the dialysis is free, there are other expenses. It is about 20-25 thousand every month which includes travel fare, fooding, sometimes lodging and medicinal cost. I would request Nepal Government to make the medicines free of cost…… P10 Patients highlighted the insecurity of living as a tenant near the hospital, with concerns about rising costs and the possibility of eviction. Patients appealed to the government or organizations to build affordable housing for vulnerable patients and even offered to contribute financially. This suggests a need for long-term housing solutions for patients who require frequent and prolonged treatment.

> ……I reside as tenant here, nearby hospital. It is costly on one hand and on other hand, the house owner could ask to leave anytime. I appeal for building for vulnerable patients like us, by government or any other organization. I am ready to assist financially from my side as well. This would aid other patients like us in future too……P11

### Quality of life of end stage kidney disease patients

The kidney disease quality of life – 36 instrument (KDQOL-36) is the tool consisting of 36 items (I) scoring from 0 to 100 for each item. Table 3 provides data for 20 patients (P1-P20), on different aspects of their quality of life, measured using the KDQOL-36 tool, which includes the following scales:

1. Symptom/Problem List: Higher scores indicate fewer symptoms and better quality of life related to symptoms of kidney disease.
2. Effects of Kidney Disease: Higher scores suggest less interference of kidney disease in daily life.
3. Burden of Kidney Disease: Higher scores indicate a lower perceived burden of the disease.
4. SF-12 Physical Health Composite: Higher scores represent better physical health and functioning.
5. SF-12 Mental Health Composite: Higher scores reflect better mental health and emotional well-being.

**Table 3.** Scales of kidney disease quality of life.

| ID | Symptom/<br>problem list | Effects of<br>kidney disease | Burden of<br>kidney disease | SF-12 Physical<br>Composite | SF-12 Mental<br>Composite |
| --- | --- | --- | --- | --- | --- |
| P1 | 64.58 | 40.63 | 25.00 | 29.34 | 54.33 |
| P2 | 43.75 | 50.00 | 12.50 | 57.14 | 44.69 |
| P3 | 47.92 | 46.88 | 18.75 | 37.11 | 49.20 |
| P4 | 89.58 | 68.75 | 43.75 | 42.61 | 55.48 |
| P5 | 39.58 | 53.13 | 0.00 | 27.36 | 46.78 |
| P6 | 56.25 | 68.75 | 68.75 | 54.16 | 34.31 |
| P7 | 31.25 | 65.63 | 12.50 | 24.96 | 47.57 |
| P8 | 68.75 | 65.63 | 6.25 | 39.01 | 38.50 |
| P9 | 60.42 | 59.38 | 18.75 | 42.49 | 41.92 |
| P10 | 43.75 | 65.63 | 0.00 | 24.81 | 48.68 |
| P11 | 97.92 | 78.13 | 56.25 | 52.66 | 54.09 |
| P12 | 81.25 | 40.63 | 6.25 | 28.10 | 42.97 |
| P13 | 72.92 | 43.75 | 25.00 | 41.80 | 48.11 |
| P14 | 70.83 | 40.63 | 6.25 | 37.29 | 51.58 |
| P15 | 58.33 | 59.38 | 0.00 | 45.93 | 23.83 |
| P16 | 50.00 | 46.88 | 6.25 | 26.19 | 43.23 |
| P17 | 64.58 | 31.25 | 0.00 | 22.43 | 49.96 |
| P18 | 58.33 | 34.38 | 25.00 | 50.31 | 49.46 |
| P19 | 85.42 | 68.75 | 37.50 | 37.38 | 54.15 |
| P20 | 77.08 | 53.13 | 25.00 | 48.22 | 33.59 |

P11 has the highest scores on several scales (Symptom/Problem List: 97.92, Effects of Kidney Disease: 78.13, Burden of Kidney Disease: 56.25), suggesting they experience fewer symptoms, less interference from kidney disease, and a lower burden. They also have strong SF-12 Physical and Mental scores, indicating relatively good overall well-being. P6 scores very high on Burden of Kidney Disease (68.75) but lower on the Mental Health Composite (34.31), which may indicate that while they feel the disease is not a heavy burden, their mental health is still affected. P17 has very low scores on the SF-12 Physical Composite (22.43) and a Burden of Kidney Disease score of 0.00, indicating significant physical limitations and a high perceived burden of kidney disease. P15 has the lowest SF-12 Mental Composite score (23.83), suggesting substantial mental health challenges, despite having moderate physical health (45.93). P4 and P11 show overall good scores across most scales, suggesting better quality of life and fewer problems related to both physical and mental health.

This data reveals a wide variation in patients’ quality of life across physical, mental, and kidney disease-specific measures. Some patients, such as P11, reported high quality of life, while others, such as P17 and P15, faced significant physical and mental health challenges.

The mean, median and standard deviation of symptom/problem list, effect of kidney disease, burden of kidney disease, SF-12 physical health composite and SF-12 mental health composite of kidney disease quality of life are illustrated in table 4.

**Table 4.** Mean, median and standard deviation of scales of kidney disease quality of life.

| Scale (number of items in scale) | Mean | Median | Standard deviation |
| --- | --- | --- | --- |
| Symptom/problem list (12) | 63.13 | 62.50 | 17.69 |
| Effects of kidney disease (8) | 54.06 | 53.13 | 13.30 |
| Burden of kidney disease (4) | 19.69 | 15.63 | 19.37 |
| SF-12 Physical Health Composite | 38.47 | 38.20 | 10.83 |
| SF-12 Mental Health Composite | 45.62 | 47.84 | 8.10 |

The relatively high mean score of 63.13 indicates that patients report a moderate level of symptoms related to kidney disease, suggesting that they experience some discomfort or challenges, but it’s not at a severely high level. The standard deviation of 17.69 suggests considerable variability in symptom experiences among patients; some may report significant symptoms, while others have fewer issues.

The mean score of 54.06 on the effects of kidney disease indicates that patients feel a moderate impact on their daily lives due to their condition. A lower standard deviation (13.30) compared to the symptom/problem list indicates that most patients have a similar perception of the effects of kidney disease, which may relate to daily activities and social interactions.

The low mean score of 19.69 suggests that, on average, patients perceive a minimal burden from their kidney disease. However, the high standard deviation of 19.37 indicates substantial variability; some patients may feel overwhelmed by their condition, while others do not perceive significant burdens. This contrast highlights the need for individualized patient support and interventions.

The mean score of 38.47 in the Physical Health Composite indicates that patients experience a lower quality of life concerning physical health. This is concerning, as it suggests that physical limitations significantly impact their daily functioning. The standard deviation of 10.83 indicates moderate variability in physical health perceptions among patients.

The mean score of 45.62 for Mental Health Composite suggests that patients’ emotional and psychological well-being is moderately affected. A standard deviation of 8.10 indicates less variability in mental health scores, meaning most patients report similar levels of mental health impact.

The overall finding suggests that kidney disease moderately impacts both physical and mental health, with significant variability in how patients experience the burden of the disease. Patients report more physical limitations compared to emotional or mental health challenges.

### Background characteristics of service providers

The dialysis section of Parbat hospital consists of seven (7) healthcare providers among which one (1) is a male Medical Officer and remaining six (6) are female nursing staffs. The service duration of dialysis service providers ranged from 3 years to 3 months while five (5) out of seven (7) of them had received kidney dialysis training. The background characteristics of the dialysis service providers is illustrated as below in the table 5.

**Table 5.** Background characteristics of service providers.

| Service Provider | Academic qualification | Designation | Sex | Kidney Dialysis |  |  |
| --- | --- | --- | --- | --- | --- | --- |
|  |  |  |  | Total experience | Experience in Parbat Hospital | Training (Yes/No) |
| SP1 | MBBS | Medical Officer | Male | 2 years 2 months | 2 years 1 month | Yes |
| SP2 | B. Sc. Nursing | Staff Nurse | Female | 3 years | 10 months | No |
| SP3 | PCL Nursing | Staff Nurse | Female | 2 years 5 months | 2 years 5 months | Yes |
| SP4 | PCL Nursing | Staff Nurse | Female | 2 years 3 months | 2 years 3 months | Yes |
| SP5 | PCL Nursing | Staff Nurse | Female | 2 years | 2 years | Yes |
| SP6 | PCL Nursing | Staff Nurse | Female | 1 year 8 months | 1 year 8 months | Yes |
| SP7 | PCL Nursing | Staff Nurse | Female | 3 months | 3 months | No |

### Experience of dialysis service providers

i. Patient personalized care and understanding patients’ behavior, Service providers expressed the feeling of being part of a family with their patients. Since dialysis patients come twice a week, healthcare providers interact with them regularly and develop a close, ongoing relationship, unlike in other wards where patients may only be seen once or for a limited time. This repeated interaction creates a bond, making the dialysis section feel more like a family environment where both patients and providers know each other well.

> ……A patient comes twice a week for dialysis, we communicate with them regularly. It is different from (other) wards, we have limited patients here and we interact with them only. It feels like living in a family……SP1

> ……Patients come twice a week, we meet them regularly. Unlike OPD and IPD, where patients stay for certain duration, may or may not meet again, we meet the patients every 4^th^ day. We are here as a family……SP2 Service providers discussed how the 4-hour dialysis sessions provide time for healthcare providers to counsel patients on essential aspects of their care, including diet, fluid restriction, hygiene, fistula care, medication, and follow-ups. They note that some patients, especially those who are health-conscious, require less frequent counseling, while others need regular reminders and guidance at every session. This personalized approach demonstrates the importance of patient education and consistent communication in managing long-term health conditions like kidney disease.

> ……During dialysis session, in the 4 hours period, we counsel the patients about diet, fluid restriction, hygiene maintenance, fistula care, medication and follow-up to almost all patients to those who are health conscious, we do not need to explain to them every time while to others, who do not follow-up regularly with the suggestions, we counsel them on every sessions……SP3 Service providers mentioned that while some patients may be uncooperative, often due to high blood pressure or emotional stress, the healthcare providers approach these situations calmly and composedly. They understand that patients might be going through physical and emotional challenges, and they make an effort to remain patient and compassionate in their interactions.

> ……There are some patients who are uncooperative. However, we take them normally. They have high blood pressure and sometimes are high tempered. We understand them and present ourselves calmly and composedly…… SP1 Service providers explained that when patients are uncooperative, the healthcare team focuses on counseling them and providing explanations about their condition. This suggests that education and understanding are key strategies for addressing patient non-cooperation, helping them manage their health more effectively.

> ……For the patients who are uncooperative, we do counselling and explanation of the condition……SP2
ii. Addressing queries and concerns of the patients, Service providers pointed out that some patients are unaware that their kidneys are completely damaged and that dialysis is necessary. They may hold onto the belief that their condition is curable, repeatedly asking when they will be cured. This highlights the importance of effective communication and education regarding the irreversible nature of kidney failure and the role of dialysis as a life-sustaining treatment.

> ……Some patients do not know the kidneys are completely damaged and dialysis is indicated, they think the condition is curable. They keep on asking when the condition will be cured……SP2 Service providers reflected that patients frequently ask questions about medications, the functioning of the dialysis machines, and the specifics surrounding kidney transplantation, including who can perform the procedure, associated costs, and success rates. This indicates a strong desire for information and understanding, demonstrating that patients are engaged and concerned about their treatment options and outcomes.

> ……Patients ask about medication, function of the equipment of the machine and transplantation – who can do transplantation, cost, success rate and so on……SP3
iii. Rewarding aspect of the job, Service providers mentioned that their formal education did not cover dialysis in depth. However, through hands-on experience and specific kidney dialysis training, they have gained significant knowledge and skills while working in the hospital. This indicates that practical, on-the-job learning plays a crucial role in building expertise in dialysis care.

> ……We haven’t learnt this much during our study (college). We were sent to kidney dialysis training as well and we are able to learn more about kidney dialysis working here……SP2 Services providers expressed gratitude for having the opportunity to participate in dialysis training. This suggests that ongoing training opportunities are valued by healthcare professionals, as they enhance their skills and improve patient care.

> ……I got opportunity of participating in dialysis training……SP3 Service providers shared their plan to work abroad as a dialysis nurse, emphasizing that the experience gained while working in the current setting has been instrumental in preparing them for future career opportunities. This reflects the importance of gaining practical experience in developing professional competencies and advancing career goals.

> ……My plan is to go abroad. I want to work as a dialysis nurse there. Working here has given me the opportunity of experience……SP5
iv. Learning acceptance and witnessing positivity, Service providers described how interacting with patients, even during the most challenging periods of their lives with kidney disease, has taught them the value of acceptance. Witnessing patients’ face their illness with resilience has been an eye-opening experience, highlighting the strength and perseverance of those undergoing long-term dialysis.

> ……Many patients have taught me acceptance, even during the difficult time of life (with kidney disease),……SP2 Service providers also noted that many patients initially feel hopeless when starting dialysis, unsure about their future. However, over time, these same patients begin to express a more positive and motivated outlook. This transformation has had a profound effect on them, teaching them that no matter how difficult a situation may seem, there is always potential for improvement and hope.

> ……Patients used to be hopeless in the beginning of dialysis regarding life. With time, patients talk about their positive motivation. This has impacted me as at the end, things can go good and we should not be hopeless……SP3
v. Availability of safety measures and protective protocols, Service providers indicated that the staff is provided with personal protective equipment (PPE), including gowns, gloves, masks, and caps, to ensure their safety during dialysis procedures. The gowns are washed and reused, demonstrating an effort to maintain hygiene through regular cleaning, while reusing materials due to practical limitations.

> ……We are provided gown, gloves, masks and cap for safety. The gown are washed regularly and reused……SP2 Service providers pointed out that while the staff has recently been provided with PPE, they are concerned about their regular exposure to blood. The team has proactively requested Hepatitis B vaccinations for additional protection, especially since there is a known Hepatitis-positive patient in the facility. This highlights the ongoing risk faced by healthcare providers and their need for comprehensive safety measures beyond just PPE.

> ……We have masks, gowns, gloves and caps recently as PPE. We have requested for Hepatitis B vaccination for safety since we are exposed to blood regularly. We have a hepatitis positive case as well here……SP3
vi. Training and professional development experience, Service providers mentioned that they have received one formal training on kidney dialysis, and there is a good balance between the training content and the actual responsibilities of the job. However, they note the absence of refresher training, which could be beneficial for updating their skills. On the positive side, technicians provide ongoing, practical guidance when new equipment and machines are installed, which helps them stay informed about technological advancements and operational procedures.

> ……I have taken one training on kidney dialysis. There is balance between the contents of training and the nature of job. We do not have refresher training. Technicians come and teach us on regular basis when new equipment and machines are installed…… SP2
vii. Reflection on job security, Service providers highlighted the lack of job security tied to the fiscal year cycle. As the end of the fiscal year approaches, they feel anxious about having only one month left to serve. They advocate for contracts to be based on work performance and the quality of service rather than influenced by external factors like power dynamics, which can create an unstable work environment.

> ……There is no security. Once Fiscal year is about to end, we feel we have only one month left to serve. The contract should be made on the basis of work performance and quality rather than power play……SP3 Service providers shared that their initial contract was for two years and is now nearing its end. While the dialysis program itself is expected to continue, there remains uncertainty about their job security. This uncertainty, despite the assurance of the program’s continuity, causes anxiety and raises concerns about the future of their employment.

> ……Initially, my contract is of two years. The two years is about to end. My appointment is from the program (kidney dialysis) and the administration has ensured the program is continuous. However, there are uncertainties regarding the job security……SP6
viii. Challenging aspect of the job Service providers expressed a mix of emotions. They feel happy to be directly involved with patients during dialysis but feel bad when complications arise that require referring patients to a higher center like Pokhara due to the absence of a nephrologist at their facility. This emphasizes the emotional toll healthcare providers face when they cannot offer complete care due to resource limitations.

> ……We are directly involved with the patients during dialysis. We feel happy and on the other hand, we feel bad when problems are encountered during dialysis and patients are referred to Pokhara (higher centre) since we do not have nephrologist here……SP1 Service providers mentioned the absence of pre-placement health check-ups, citing an instance where a dialysis patient was later discovered to be HIV-positive and had syphilis. While the situation was handled by conducting the necessary tests, it reveals a potential health risk to healthcare providers who may unknowingly be exposed to infectious diseases.

> ……There was no pre-placement health check-up. Once a dialysis patient was a HIV positive patient and also had syphilis, we informed the hospital authority for test and respective serological tests were done…… SP2 Service providers described the difficulties of managing sudden complications, such as when a patient’s blood pressure dropped during dialysis and did not stabilize despite administering medications and stopping the treatment. This illustrates the critical nature of dialysis care and the challenges faced when complications arise that are beyond their control.

> ……If complications arise in patients, I find difficulty. Once a patient’s blood pressure declined suddenly, we gave medicines, stopped the dialysis, however, the blood pressure did not rise to normal……SP2 Service providers shared that the nature of their job feels repetitive and monotonous, with little opportunity to learn something new due to the daily routine being the same. This reflects the mental fatigue that can come with repetitive tasks in healthcare, especially when the responsibilities do not vary.

> ……The nature of job is monotonous. There is no new thing to learn because the responsibility is same daily……SP3 Service providers also expressed concern over the potential exposure to blood, chemicals, and contagious diseases like tuberculosis (TB) in their work environment. This highlights the occupational hazards healthcare providers face while caring for patients, especially in settings with inadequate protective measures.

> ……We are exposed to bloods and chemical. We have TB cases. We might get contaminated……SP5 Service providers noted that technical and logistical issues, such as machine malfunctions, power cuts, and water shortages, frequently disrupt dialysis sessions. These operational challenges not only create stress for the healthcare providers but also impact the quality of care for the patients, making the experience more difficult for everyone involved.

> ……Sometimes the dialysis machines stop working, we have to pause the process, after the dialysis has started, example after one hour, sometimes, there is power-cut, or shortage of water. These things make me feel bad for the patients……SP6
ix. Further expectations and requests. Service providers discussed the challenges faced due to the lack of a nephrologist and the need to refer patients to facilities in Baglung (neighboring district) or Pokhara (Province headquarter) for complications. They advocate for the establishment of an ICU ward at their hospital, which would enable them to provide better care for patients and reduce the need for referrals.

> ……We do not have a nephrologist doctor here. We have only once taken the kidney dialysis training, other than that, we would also like to get involved in aspects to upgrade our knowledge. We have to refer patients to either Baglung or Pokhara in case of difficulty to patients, if an ICU ward would be established, we could take care of patients by ourselves……SP1

> ……Our contract is temporary, of around 10 months or 11 months. My contract ends this Fiscal year. I want the hospital committee to extend the contract……SP2 Service providers reflected on the limited training received, mentioning that they have only undergone one kidney dialysis training without any refresher courses. The lack of ongoing education prevents them from learning new techniques or updates in the field, which could enhance their skills and service delivery.

> ……We have less number, basically one training for kidney dialysis. We do not get any refreshers’ training. We do not get to learn about new things……SP4 Service providers emphasized the absence of basic life support training among the staff, which they believe is essential for dialysis service providers. Such training could be critical in saving lives during emergencies, indicating a significant gap in their preparedness for acute situations.

> ……We do not have basic life support training, none of us. This is a need to dialysis service provider because it could save life in emergencies……SP5 Service providers pointed out that having a nephrologist available would greatly assist in enhancing their knowledge and providing better guidance for patient care. This underscores the need for specialized medical expertise to support both staff and patients effectively.

> ……Availability of nephrologist would assist in upgrading our knowledge. We would also have better guidance for service delivery……SP6 Service providers raised concerns about the temporary nature of their contracts, typically lasting around 10 to 11 months. With the end of the fiscal year approaching, they express a desire for the hospital committee to extend their contracts, reflecting the insecurity that comes with temporary employment in healthcare.

### Job satisfaction of service providers

Table 6 demonstrates dialysis service providers’ responses over every thirty-six (36) items of Job Satisfaction Survey (JSS). For the 36-item total where possible scores range from 36 to 216, the ranges are 36 to 108 for dissatisfaction, 144 to 216 for satisfaction, and between 108 and 144 for ambivalent. Out of seven (7) kidney dialysis service providers, all of them were ambivalent (neither satisfied nor dissatisfied) with their job where the score ranged from 115 to 135 as shown in table 6.

**Table 6.** Total JSS score and job satisfaction level of dialysis service providers.

| **Service Provider** | **Total JSS Score** | **Job Satisfaction** |
| --- | --- | --- |
| SP1 | 133 | Ambivalent |
| SP2 | 131 | Ambivalent |
| SP3 | 135 | Ambivalent |
| SP4 | 128 | Ambivalent |
| SP5 | 132 | Ambivalent |
| SP6 | 115 | Ambivalent |
| SP7 | 135 | Ambivalent |

## Discussion

The end-stage kidney disease patients visiting Parbat hospital for kidney dialysis service in this study reported wide-range of experience for those nine themes were extracted; (i) perception on end-stage kidney disease, (ii) perception on free kidney dialysis service at the hospital, (iii) administrative process for free dialysis service, (iv) feasibility with the schedules of dialysis session, (v) quality of care and response of service providers (vi) duration of dialysis session (vii) desires and challenges towards kidney transplantation, (viii) on recommending the dialysis site and (ix) further expectations for improvement.

In this study, end-stage kidney disease patients illustrated the emotional struggle and the adjustment to life with kidney disease. They highlight both the frustrations of living with physical limitations and the hope that medical treatments like dialysis can provide in prolonging life. These experiences are in line with the findings of studies conducted among dialysis patients in Nepal and Uganda.(14,15)

The dialysis patients expressed profound difference that free dialysis services and access to medications have made in their lives, emphasizing both the emotional and financial relief they get. Overall, the administrative side of accessing free dialysis appeared smooth in this study according to the patients’ reflection. The study also highlighted the variability in patient experiences based on individual circumstances, such as proximity to the hospital, ability to accommodate different schedules, and personal resources for handling extended stays. The study found that the dialysis patients have diverse experiences during dialysis session, ranging from boredom and restlessness to positive feelings about the treatment process.

Regarding the quality of care of the dialysis service, the patients in this study reflected general satisfaction with the dialysis services at Parbat Hospital, particularly regarding timeliness, cleanliness and the caring nature of the staff. The behavior and communication of the nursing staff were found to play a crucial role in enhancing patient satisfaction at the hospital, as patients feel their needs and concerns are properly addressed with kindness and professionalism. In a study among dialysis patients in Ethiopia, on a similar note, participants described that they had a good relationship with the dialysis center staff and that they were receiving kind care.(16)

With regard to kidney transplantation, the end-stage kidney patients emphasized the need for legal reforms in kidney transplantation to widen donor eligibility and address the financial barriers that prevent patients from accessing life-saving transplants. In a similar study conducted in Ethiopia, lack of a donor and sponsor were noted as barrier to having a kidney transplant.(16)

The dialysis patients in this study showed the overall satisfaction with Parbat Hospital’s dialysis services but also pointed out a critical gap in specialized care, particularly the need for a nephrologist. While many patients are happy to recommend the hospital, addressing this gap could improve the service further. The patients in this study also expressed the need for financial support for medications to better support chronic kidney patients and reduce their risks and financial stress.

The study found out, regarding quality of life of end-stage kidney disease patients, that kidney disease moderately impacts both physical and mental health, with significant variability in how patients experience the burden of the disease. Patients reported more physical limitations compared to emotional or mental health challenges. Several studies have similar findings with compromised health related quality of life among end stage renal disease patients and lower values of quality of life parameters in dialysis patients while some studies demonstrated no significant changes in quality of life of end stage kidney disease patients even when following-up.(17–19)

The service providers shared a wide range of experiences on delivering the kidney dialysis services for those nine themes were extracted; (i) patient personalized care and understanding patients’ behavior (ii) addressing queries and concerns of the patients (iii) rewarding aspect of the job (iv) learning acceptance and witnessing positivity (v) availability of safety measures and protective protocols (vi) training and professional development experience (vii) reflection on job security (viii) challenging aspect of the job (ix) further expectations and requests.

The dialysis service providers shared the unique continuity of care in dialysis, where repeated patient-provider interactions foster a supportive, family-like atmosphere and allow for ongoing patient education and personalized attention. They also highlighted the importance of patience, empathy, and communication in managing difficult interactions with dialysis patients, acknowledging the emotional and physical burdens these patients may be experiencing.

The dialysis service providers, doctor and nurses in this study were found emphasizing the value of hands-on training and experience in the field of dialysis, as well as the professional growth and aspirations of healthcare providers working in this area.

The service providers of the study highlighted the need for improved resources, better health and safety measures, and support systems to ensure that both patients and healthcare staff have a safer, more effective environment.

In this study, regarding the safety measures for service providers, the participants highlighted the current measures in place to protect healthcare workers in dialysis sections, while also emphasized the need for further protective steps, such as vaccinations, to safeguard their health in a high-risk environment.

While initial training is adequate, according to the dialysis service providers, there is a need for more structured, ongoing professional development through refresher courses to ensure that healthcare providers remain up-to-date with best practices in kidney dialysis care. Regular technical guidance is helpful but may not fully substitute for comprehensive training updates.

In this study, the dialysis service providers, particularly, nurses expressed the emotional and professional strain that job insecurity places on healthcare workers. Even when programs are ongoing, the lack of stable, long-term contracts leads to uncertainty and stress, impacting both job satisfaction and personal well-being. The healthcare providers call for a more stable and performance-based employment system that ensures security and rewards their dedication to the job.

The service providers in this study expressed the urgent need for enhanced training, job security, specialized medical support, and improved facilities in the dialysis section. Addressing these issues would not only benefit the healthcare providers but also significantly improve the quality of care provided to patients undergoing dialysis. Studies among nurses in Nepal have found continuing professional development of nurses as the major needs for quality of practice and skills.(20)

This study revealed that doctor and nurses who are the kidney dialysis service providers at Parbat Hospital were ambivalent, neither satisfied nor dissatisfied with their job under the job satisfaction survey tool. In contrary, studies revealed majority of the service providers were overall satisfied with their jobs.(7,9) However, dissatisfaction with certain areas of the job was also observed in previous studies.(21)

## Conclusion

This study sheds light on the diverse experiences of both end-stage kidney disease (ESKD) patients and service providers at Parbat Hospital’s dialysis section, offering a comprehensive view of the challenges and successes within this setting. For patients, the free dialysis service is a significant relief, particularly in reducing the financial burden of their long-term care. However, they reported varied experiences regarding the convenience of dialysis schedules, administrative processes, and the duration of sessions. Many expressed the desire for improved access to kidney transplantation and faced emotional and physical challenges in managing their condition. The high variability in the Burden of Kidney Disease and relatively low scores in the Physical Health Composite suggest that addressing physical health challenges and reducing the burden of the disease should be key areas of focus for improving the overall quality of life for these patients.

From the perspective of service providers, the study reveals both the rewarding and challenging aspects of their work. Providers found satisfaction in helping patients and witnessing their resilience, but they also noted the monotony of the job, lack of nephrology specialists, and limited professional development opportunities. Concerns about job security, the absence of essential resources such as specialized medical support, and safety measures, particularly regarding exposure to health risks, were also prevalent. The ambivalent satisfaction level of service providers also adds for further addressing the concerns.

Despite these challenges, both patients and providers showed optimism for future improvements, including better training programs, enhanced infrastructure, and more robust medical support systems. Addressing these issues could not only improve the quality of life for patients but also enhance job satisfaction and performance among healthcare workers. By incorporating patient feedback and service provider input, hospital administration and policymakers can develop more effective strategies to strengthen the dialysis services offered at Parbat Hospital.

## Data Availability

All data produced in the present study are available upon reasonable request to the authors

## Acknowledgement

I would like to extend my thanks to Parbat Hospital for granting permission to conduct this study. My special thanks to all the end kidney dialysis patients and service providers of the dialysis section who provided their valuable time and information.

